# Student Health Initiatives for Enhanced Disease Surveillance (SHIEDS) in Ghana

**DOI:** 10.1101/2025.08.03.25332924

**Authors:** Doreen Tachie-Donkor, Rosina Boateng Osei, Hillary Naki Debrah, Yvonne Osei Adobea, Emmanuel Osei Sackey-Aidoo, Janice Anum, Paul Kwadwo Addo, Charles A. Narh

## Abstract

**Background:** Student Health Initiatives for Enhanced Disease Surveillance (SHIEDS) is a student-driven program that aims to strengthen infectious disease surveillance and enhance healthy lifestyles within university communities in Ghana. This study aimed to assess SHIEDS feasibility and implementation at the Kwame Nkrumah University of Science and Technology (KNUST), Kumasi, Ghana.

**Methods:** Between 29^th^ June and 6^th^ July 2024, educational campaigns were conducted, through radio and social media, to raise awareness about sexually transmitted infections (STIs) among the student population. These campaigns ended with free screening for two STIs: HIV & Hepatitis B. Participants provided verbal feedback that were reviewed and included in a recommendation report for the KNUST administration. Positive cases were offered counselling and referred for confirmatory testing at the University Hospital, KNUST, Ghana.

**Results:** The SHIEDS awareness campaigns reached more than 20,000 people through social media and the radio outreach benefitted over 3,000 students, with 4 and 5-star ratings for overall program delivery and media campaigns, respectively. A total of 228 students, with mean age of 23 years (range of 18-29) consented to screen for STI by rapid diagnostic testing. The combined STI positivity rate was 0.87%, with rates of 1.01% for hepatitis B and 0.77% for HIV detection among males and females, respectively; all being self-reported old cases on active treatment.

**Conclusion:** Review of student feedback recommended screening for other STIs including gonorrhoea, syphilis and chlamydia, and instituting SHIEDS as an annual event in the university calendar. Feasibility studies in other universities will inform program standardization and implementation across Ghana. Our findings indicated a healthy student community, which could serve as reference for future SHIEDS programs in KNUST.

## Introduction

Infectious diseases remain among the leading causes of mortality in Africa (1). While significant progress has been made in combating certain conditions, infectious diseases remain major contributors to the continent’s disease burden; the five leading infectious disease killers – acute respiratory infections, human immunodeficiency virus (HIV), diarrhoea, malaria, and tuberculosis account for nearly 80% of the total infectious disease burden. These diseases claim over 6 million lives annually (2). Notably, sexually transmitted infections (STIs) pose health threats including acute illnesses, infertility and high mortality rates among young people (3).

STIs remain a significant public health challenge in Africa due to their high prevalence and serious health consequences. A Kenyan study involving 400 females aged 16–20, nearly three-quarters reported engaging in sexual activity, of which more than 55% had at least one STI (4). Similarly, a study in South Africa, found a high prevalence of STIs among young people, with HIV prevalence rates of 5.6% in men and 19% in women aged 15-24 years (5). In Africa, STIs result in tremendous poor health outcomes including increased healthcare costs and reduced productivity (6). Thus, public health control interventions including health education and screening of vulnerable population are crucial to reducing STI burden in Africa.

Globally, HIV is a leading STI causing death among women aged 20-40 years (7). Most HIV infections remain asymptomatic, often going undetected under national syndromic management guidelines (8). This underscores the importance of STI surveillance among vulnerable populations to enhance control programs. In Ghana, for example, HIV has evolved from a health issue to one affecting all socio-economic aspects of life (9). Public Health strategies, such as education and early screening exercises are crucial for raising awareness and reducing the impact of HIV (10).

Other STIs including hepatitis B virus (HBV) infection, affects ∼300 million people globally, and is the leading cause of cirrhosis and liver cancer (11). Major medical complications also include acute flares and extrahepatic manifestations. HBV remains largely underdiagnosed in Africa and effective measures that can prevent infection and disease progression are underutilized (11); HBV patients experience stigma which can discourage them from seeking care. Despite the goal of the World Health Organization to eliminate viral hepatitis as a public health problem by 2030, the annual global deaths from HBV are projected to increase by 39% in 2030 if proper control efforts including diagnosis are not scaled-up (12).

Among the populations at risk for STIs, the youth, 15-35 years age group, account for a substantial portion of the global infectious disease burden (13). In 2019, an estimated 30 million youth died of infectious diseases, representing 57.3% of the global communicable disease burden. Additionally, this age group lost over 30 million years of healthy life due to disability, with cumulative impact of 288.4 million disability-adjusted life-years (DALYs) (14). These statistics highlight the urgent need for targeted interventions that focus on the youth, including comprehensive sexual health education, early screening, and youth-friendly healthcare services to reduce the long-term impacts of STIs.

This Student Health Initiatives for Enhanced Disease Surveillance (SHIEDS) exemplifies an innovative, student-led program to raise awareness about infectious diseases and to promote healthy lifestyle choices that impact health of student communities in Ghana. This report discusses SHIEDS feasibility and implementation at the Kwame Nkrumah University of Science and Technology (KNUST) campus in Kumasi, Ghana.

## Methods

### Catchment Area

SHIEDS was conducted, between 29^th^ June and 6^th^ July 2024, on the main campus of KNUST, located in Kumasi, within the Oforikrom Municipal Assembly of the Ashanti Region, Ghana (Figure 1A). STI awareness campaigns covered five communities: Ayeduase, Kotei, Ayigya, Kentinkorono and Boadi. These communities are areas of major socioeconomic activity, with residential facilities for the university community. In 2021, the municipality had a population of 213,000, with nearly equal proportions of males and females. It had a youthful population, with 64.5% being in the age group 10-39 years, and a literacy rate of 90% (15).

**Figure 1.**
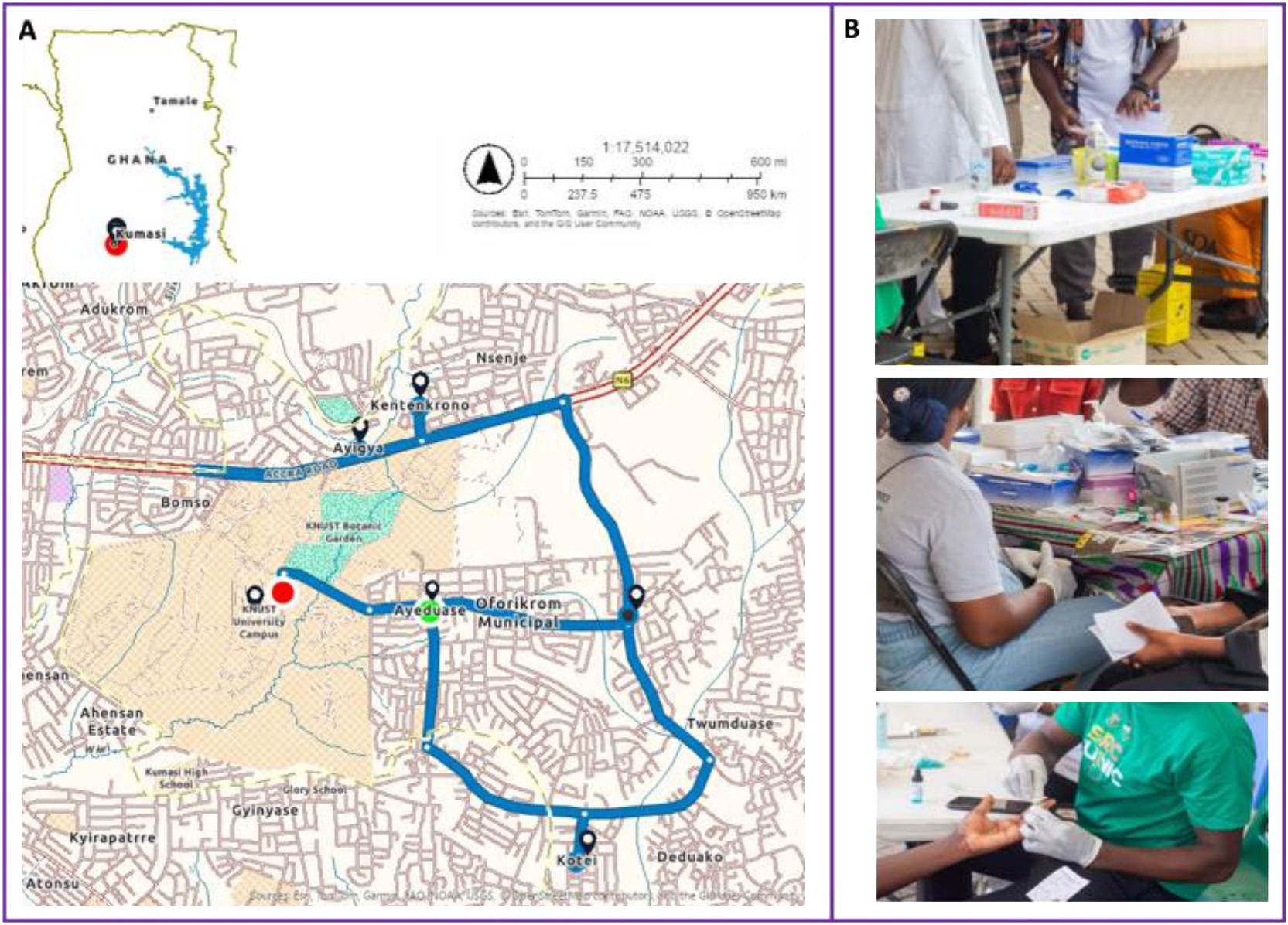
SHIEDS catchment areas and screening activities. **A.** Map of Ghana (top) and extended map of the Oforikrom Municipal including the SHIEDS communities. The catchment areas – Kentenkrono, Ayigya, Ayeduase, Oforikrom, Twumduase and Kotei are well connected by major road networks (blue), located within Oforikrom municipality, Ashanti Region. The municipal comprises urban and peri-urban settlements including student residential facilities. Key landmarks within the catchment area include KNUST university campus (red), which serves as a central reference point for faculty and student activities. **B**. Photos of SHIEDS activities including booths for participant data collection and counselling and booths for finger-prick blood collection and STI testing.

### Media Outreach

To raise awareness and promote participation among the student community, the SHIEDS program was advertised on social media – Twitter/X and LinkedIn and through interviews on national radio – KNUST’s campus radio and Focus FM. These awareness programs were projected to reach the estimated 85,000 student population. Therefore, the messaging focused on the importance of STI screening, HIV and hepatitis B (HBV), among sexually active adults; it was also disseminated through KNUST and student Influencer X pages, over 60k followers.

### Health Information/Education and Awareness

On the morning of the health screening, participants were educated about STIs – modes of transmission, treatments, control and prevention. Additionally, infographic posters were mounted at vantage points around campus to disseminate facts about STIs. Free condoms were distributed to eligible adults aged 18 and above, and private consultations/counselling were offered to participants by medical staff from the Health Directorate.

At the start of the HIV screening, the majority of the participants seemed nervous, likely due to the stigma associated with being identified as ‘HIV patient’ in some cultural settings. Therefore, we reassured them through one-on-one confidential counselling, including benefits of knowing one’s ‘HIV status’, and available treatments options. We provided information about certified hospitals where suspected cases could go for confirmatory test and access treatment.

### Health Screening

The health screening activities were conducted on campus (Figure 1B), and targeted students 18 years and above. Four designated booths (Figure 1B) were set up – Infectious Diseases checks – HIV (First Response HIV 1-2. O Card Test, Premier Medical Corporation Private Limited), malaria (SD Biosensor Malaria Pf/Pan Ag) and HBV (Advanced Quality One-Step Multi-HBV Test Kit). General health checks included Body Mass Index (BMI), blood pressure (Omron M1 Basic Automatic Blood Pressure Monitor, Omron Corporation) and body temperature (Omron digital thermometer, Omron Corporation). In collaboration with the School of Dentistry and the Department of Optometry at KNUST, we also provided dental and eye screenings (not included in this report).

## Results and Discussion

### Media Education and Awareness

The media campaign reached an estimated 20,000-30,000 views across social media. The information session (flyers and posters) and radio interviews reached and benefitted over 3,000 students, with 4 and 5-star ratings for overall program delivery and media campaigns, respectively. Based on verbal responses that we received during the information sessions, it seemed most students were aware of STIs including transmission, prevention and treatment options; example, nearly all participants had heard of and/or watched adverts about antiretroviral therapies for HIV and vaccination for HBV. While we did not specifically ask questions on sexual behaviour, most participants self-reported knowing about safe sex practices including the use of condoms.

### Health Screening Outcomes

A total of 228 students, with mean age of 23 years (range 18-29), including 43.4% and 56.6% identifying as male and female, respectively, participated in the screening program. All participants provided informed verbal consent prior to STI (HIV and HBV) testing, with option to also screen for malaria.

Test results

- HIV: 1/228; positivity rate of 0.43% (overall) and 0.77% (females).
- HBV: 1/228; positivity rate of 0.43% (overall) and 1.01% (males)
- Malaria: No positive cases.

STI prevalence was 0.87%, all being self-reported old cases on active treatment. The study rates were lower than the national average reported for HIV (1.7%) and HBV (8.3%) among adults in a 2019-2020 study (16, 17). Access to care and effective disease control efforts within the KNUST community will be seminal for future programs. It is possible that students who participated in our program already knew their STI status, which needs to be encouraged and promoted. Malaria co-infections can complicate STI clinical disease (18). In Ghana, most malaria infections in adults are asymptomatic (19), with community screening program reporting 6% prevalence in 2024 (20). No malaria case was identified in this program. Our findings portray a healthy student community, which serves as a reference for future screening programs in KNUST.

### Recommendations for Student Wellbeing

Our review of student feedback recommended screening for other STIs including gonorrhoea, syphilis and chlamydia, and instituting SHIEDS as an annual event in the university calendar. Feasibility studies in other universities will inform standardization and implementation of SHIEDS across the country. Future programs are encouraged to focus on student wellbeing and include Basic Life Support (BLS) training such as Cardiopulmonary Resuscitation (CPR), supported with disability aids and mental health services. Digital health solutions such as tele-counseling/consultation, with online appointment apps can help improve service accessibility and efficiency. Food services on campus are vital to student health and therefore need institutional regulation in partnership with vendors.

## Conclusion

The SHIEDS program at KNUST was well accepted by students and has prospects to improve community health through disease surveillance and healthy lifestyle promotions. By actively involving students, the program fostered a sense of responsibility and accountability among students towards raising awareness about STIs control and prevention in Ghana.

## Acknowledgement

This project was carried out by the Health Committee of the Student Representative Counsel at KNUST, with financial and logistic support from the Directorate of Students Affairs (DoSA), KNUST and the University Hospital, Oforikrom Municipal Assembly - Health Directorate, Ashanti Region, Ghana. The SRC Health Committee is gratitude for the diverse support it received from Prof. Wilson Agyare (Director, Directorate of Students Affairs, DoS - KNUST), Dr. Osei Kwaku Wusu-Ansah (Director of Health Services, KNUST), Ms. Yvonne Osei Adobea (KNUST SRC President – 2023/2024 Academic Year), and thank Dr Kwame Akrasi, Mr. Kwabena Fosu, Edmund Gali, Jessica Boampong and Julie Afoakwa for their technical support/advice.

## Author Contributions

Conceptualisation: DTD & CAN. Investigation and methods: All authors. Data curation: DTD, CAN & RBO. Formal analysis: DTD & CAN. Supervision: CAN & PKD. Manuscript draft and formatting: DTD & CAN. Review and editing: CAN, KPD, HND & RBO. All authors have read and approved the manuscript for publication.

## Institutional Review Board Statement

The SHIEDS program including media education and health screening at KNUST received approval from both the University’s Director of Student Affairs and the Director of University Health Services. The program also received technical and logistics support from the Oforikrom Municipal Health Directorate, Oforikrom Municipal.

## Informed Consent Statement

Study participants provided informed verbal consent.

## Data Availability Statement

Provided in the manuscript

